# SARS-CoV-2 post-vaccine surveillance studies in Australian children and adults with cancer: SerOzNET Statistical Analysis Plan

**DOI:** 10.1101/2023.02.26.23286261

**Authors:** Amy Body, Catherine Martin, Lucy Busija, Luxi Lal, Elizabeth Ahern, Raina C MacIntyre, Eva Segelov

## Abstract

COVID-19 disease is associated with higher morbidity and mortality in cancer patients. Our study aimed to characterize the optimal strategy to improve vaccine induced protection against COVID-19 in children and adolescents with cancer. Results from The SerOzNET study will contribute comprehensive data on serology, cellular immune correlates from functional T-cell assays, quality of life data, and associated toxicity in relation to COVID-19 vaccination in children and adults with cancer.

In this plan, we describe the statistics that will be used to report results of the SerOzNET study. SerOzNET examines COVID-19 vaccine response in children and adolescents with cancer.

We have no conflicts of interest to disclose.

## Study synopsis

### Study design

SerOzNET is a prospective observational cohort study, where the response to COVID-19 vaccination is measured at multiple time points amongst different cohorts of patients with cancer.

### Study aims

1. To determine the seroconversion rate in different cancer cohorts in children and adults
2. To determine if the seroconversion rate differs among the different cancer cohorts in children and adults
3. To determine the associations with seroconversion
4. To determine relationship between seroconversion and **T** cell response

Further details of the trial design are available in the published protocol paper. (1)

During study enrolment (mid-2021 to mid-2022), healthy adults and older adolescents (16+) in Australia were recommended to receive a minimum of 3 COVID-19 vaccine doses. Healthy children aged 5-15 were recommended to receive 2 COVID-19 vaccine doses. Of note, children aged <12 years receive a reduced dose of vaccine compared to adolescents and adults. This dose is not adjusted according to patient weight.

Since inception of the study, additional doses of vaccine have been made available to vulnerable groups including patients with cancer, including additional doses in the primary course and a “winter booster” dose. The protocol was amended after these doses were added to add observational time points after each dose.

### Study design

All patients have a current or previous cancer diagnosis. Two groups; adults (20 years and above) and children/adolescents (5-19 years) will be analysed separately.

Both groups are split into 5 cohorts:

1. Haematological cancer
2. Solid cancer on chemotherapy (within 28 days of dose 1)
3. Solid cancer on immunotherapy
4. Solid cancer on targeted or hormonal therapy
5. Solid cancer completed chemotherapy 29-365 days before dose 1 The children and adolescent group has an additional cohort
6. patients who received systemic treatment more than 365 days ago, or never received systemic treatment.

## Endpoint Definitions

### Primary Endpoint

Proportion of patients in each cohort who develop a detectable neutralising antibody response (defined as a neutralisation titre 1:20) post COVID-19 vaccination one month after completion of the standard course (2 doses for children/adolescents, 3 doses for adults).

### Secondary Endpoints

1. Comparison of seroconversion rate between cancer cohorts and a healthy control group
2. Comparison of seroconversion rate between cancer cohorts, with the haematology cancer cohort group as the reference group
3. Proportion of patients in each cohort who develop a protective neutralizing antibody response at 1 month and 3 months following each vaccination dose subsequent to the initial dose
4. Proportion of patients in each cohort who develop a detectable T cell response at 1 month and 3 months following each vaccination dose subsequent to the initial dose
5. Comparison of spike protein lgG geometric mean titres (GMT) between cancer cohorts at each time point
6. Change in geometric mean lgG antibody titre at serial time points

### Exploratory endpoints

1. Examine the pattern of serological response
2. Examine the pattern of T cell response
3. Examine the effect of epigenetic T cell exhaustion signature on serological and T cell response
4. Examine rate of seroconversion in each cancer cohort against a healthy control population
5. Examine the effect of haematological treatments of concern on outcome (haematological cancer population only)
  a. Purine analogues, proteasome inhibitors, R-CHOP, anti CD20, Anti Bcl-2, BTK inhibitors, autologous stem cell transplant, allogenic stem cell transplant
6. Examine the effect of previous stem cell transplant on outcome (haematological cancer population only)
7. Examine the effect of change in treatment or cancer status after completion of the routine course of vaccination (3 doses adults, 2 doses children) on the response to subsequent booster doses

### Safety Endpoints

- Patient reported adverse events in each cohort (Patient Reported Toxicity-CTCAE)
- Medically determined adverse events at each time point
- Patient and/or clinician reported COVID-19 infection

#### Sample size

The sample size of 100 per cohort will have over 80% power(of 0.80) to detect a decrease of 10% seroconversion rate in any cancer cohort (compared with assumed non-cancer population incidence of 95%) with 95% confidence (a of 0.05). (1)

## STATISTICAL ANALYSES

### General principles

The analysis and reporting of the results will follow the STROBE guidelines for observational studies. (2)

Baseline characteristics will be tabulated by using appropriate summary statistics. Interpretation of the primary endpoint will be based on confidence intervals. For secondary and tertiary endpoints two-tailed P-values will be reported in addition to confidence intervals. A nominal 5% significance level will be employed.

#### Analysis Populations

##### Primary and Secondary outcome population

The primary, secondary and exploratory outcome populations will consist of all patients who consented to the study. Patients who withdraw consent after receiving the first dose will have their data used, up until the time of withdrawal unless the person specifically requested all their data to be withdrawn. Numbers and reasons for the missing primary outcome will be tabulated for each cohort (withdrew, lost to followup, deceased etc).

### Primary endpoint analysis - seroconversion rate (detectable neutralising antibody response one month after completion of the standard course)

The seroconversion proportion and its exact binomial two-sided 95% confidence interval (Cl) will be determined for each cancer cohort. The one-month definition will include neutralising antibody response result taken 4 weeks (−1 week /+2 weeks) post final vaccination of the standard course.

If there is >5% and < 40% missing primary outcome the missing seroconversion data will have characteristics compared to those who have the measurement present and there is evidence of missing seroconversion data being dependent on the characteristics then a sensitivity analyses for the impact of missing primary outcome will involve multiple imputation with chained equations with 20 imputations, separately for each cancer cohort, incorporating information from non-missing baseline and post-baseline variables in the imputation models. If the proportions of missing data is very large(> 40%) on important variables, then trial results may only be considered as hypothesis generating results. (3)

### Secondary endpoint analyses

Secondary endpoints will be compared across the different cohorts.

Binary endpoints (Secondary# 1 [S1], S2 and S3) will use log-binomial regression to estimate risk difference and risk ratios together with their 95% Cls, or exact logistic regression to approximate these values if the number of events in either arm is fewer than 5.

Continuous endpoint S4 will be assessed and if reasonably symmetric distribution linear regression with robust standard errors will be used to estimate differences in means with 95% Cls, with adjustment for the baseline value of the endpoint if available. If heavily skewed will be summarised as medians and interquartile ranges, with estimated differences between medians and their 95% Cls computed via quantile regression adjusting for baseline values if available.

The spike protein lgG geometric mean titres (GMT) will be calculated for each cancer cohort at each time point. The 95% Cl of the geometric means will be calculated with the Student’s t distribution on log-transformed data.

Log-binomial analysis will be undertaken to adjust seroconversion rate between cancer cohorts adjusting for age, sex, steroid use, ECOG, BMI, and any other potential confounder found to be imbalanced between the cancer cohort groups. Should the log-binomial model fail to converge, modified Poisson regression with robust standard errors will be used.

### Subgroup analyses

Proportion of seroconversion will be tabulated overall and by each cancer cohort for the following sub-groups:

1. Age will be categorised into 20-40, 40-50, 60-70 and 70+ years.
2. Steroid use -defined as nil, low or high dose-treatment at the time of the first vaccine dose
  1. No steroid: responses “nil” or “replacement dose only”
  2. Low “Yes chemo day only”, Yes day prior and day of chemo”, “Yes Day of and 1-3 days post chemo”
  3. High “Yes treatment dose daily” “Yes treatment dose given for haematological malignancy”
3. BMI categories defined as <18.5, 18.5 to 24.9, 25-29.9, 30+
4. ECOG categ ories defined as 0, where all categories are 0 or >0 for any ECOG category recorded as >0.
5. Patients with previous COVID infection at baseline

To determine if the there is a difference behaviour between cancer cohort and subgroup an interaction term will be included in the log-binomial model between subgroup and cancer cohort.

Additionally, a log-binary analysis will be undertaken in all participants which will include the cancer cohort, age (continuous), BMI (continuous), steroid use (binary), ECOG (binary), previous COVID infection (binary) and any other potential confounder found to be imbalanced between the cancer cohort groups. Estimated proportions will be calculated for each cancer cohort with continuous values centred around their mean.

### Exploratory endpoint analyses

Exploratory outcomes one to three will have the pattern of response graphed across time overall and by cancer cohort.

For exploratory outcome four the proportion of seroconversion for healthy adult controls will be calculated and compared to each cancer cohort separately using log-binomial regression. Due to the expected small numbers of healthy controls the only adjustment that will occur will be for age and possibly gender.

For exploratory outcomes five and six, proportion of responders by each treatment of concern will be reported and compared using log-binomial regression. For exploratory outcome seven, patient response post completion of the standard course (3 doses adults, 2 doses children) will be compared to their response to subsequent booster doses.

For adults:

- Effect of ceasing systemic therapy (chemotherapy, immunotherapy, hormone therapy or targeted therapy) will be examined
- Effect of change in systemic therapy or initiation of new systemic therapy will be examined
- Compared to patients who had no change in treatment

For children: see section on children/adolescent analysis

For binary outcomes log-binomial regression will be undertaken, if numbers are < 5 in a cell exact logistic regression will be undertaken.

### Characteristics and their definitions-see table1

#### Safety endpoint analyses

Safety endpoints will be tabulated by treatment group without statistical tests or confidence intervals.

**Table 1:**
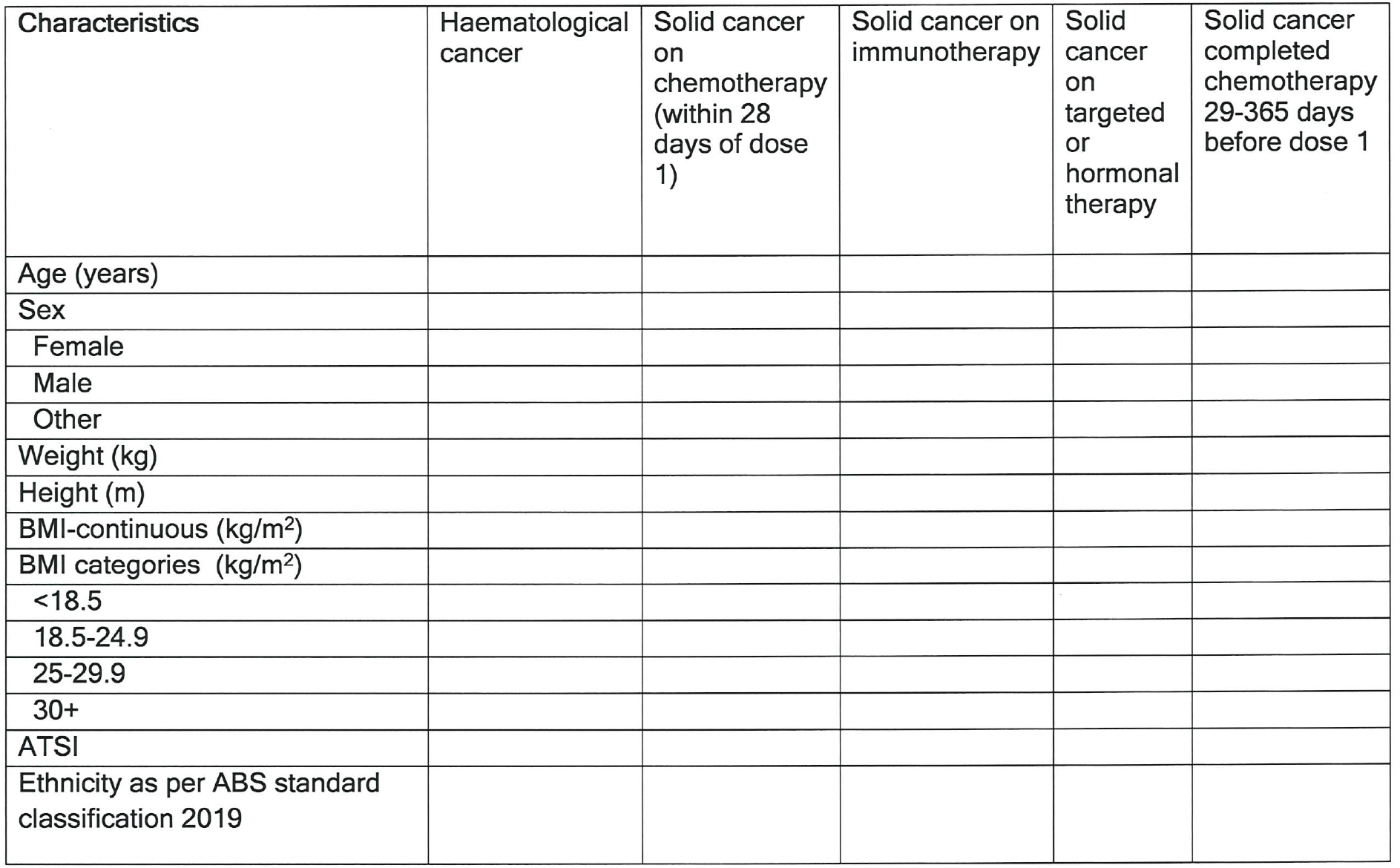
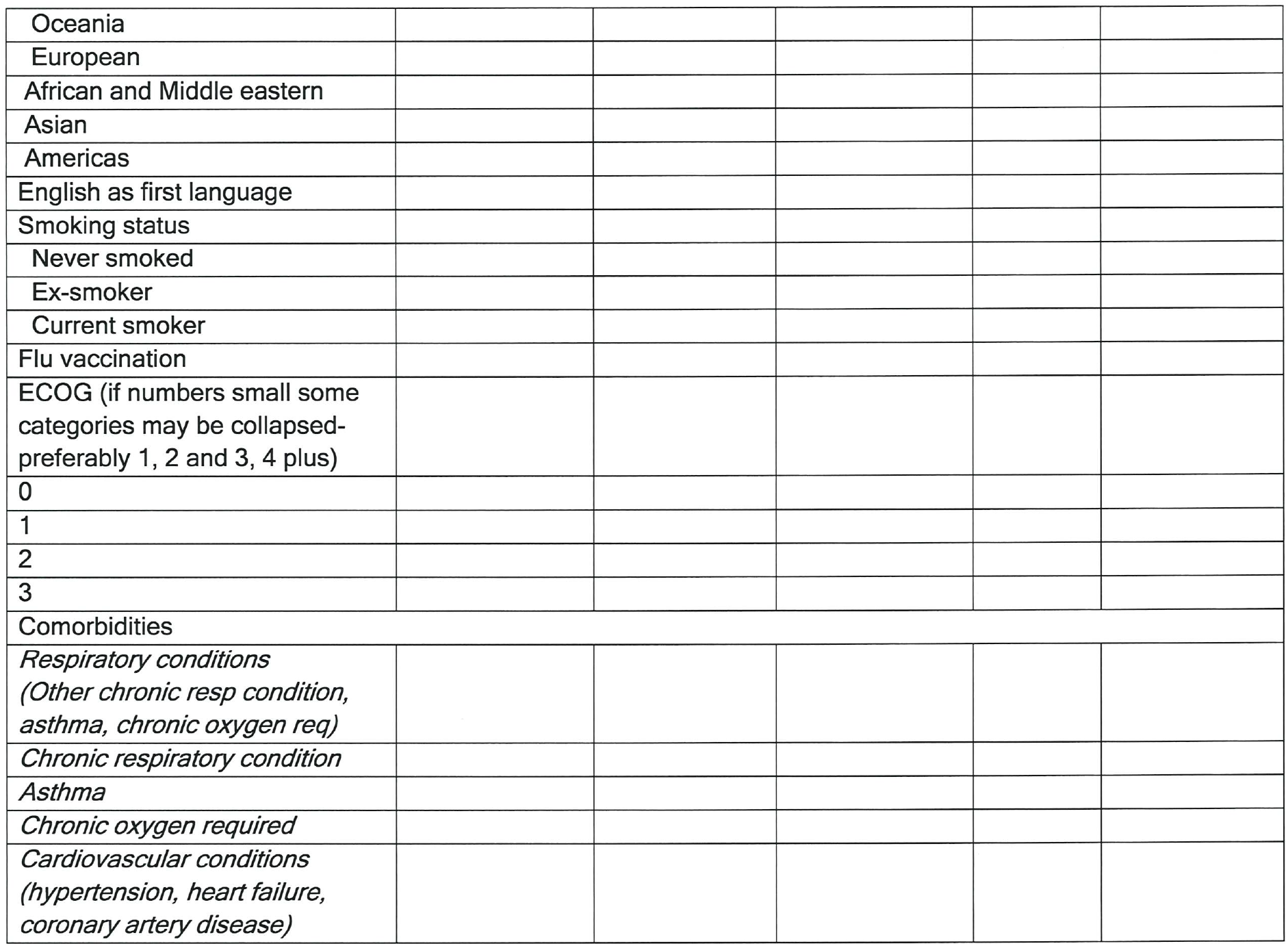
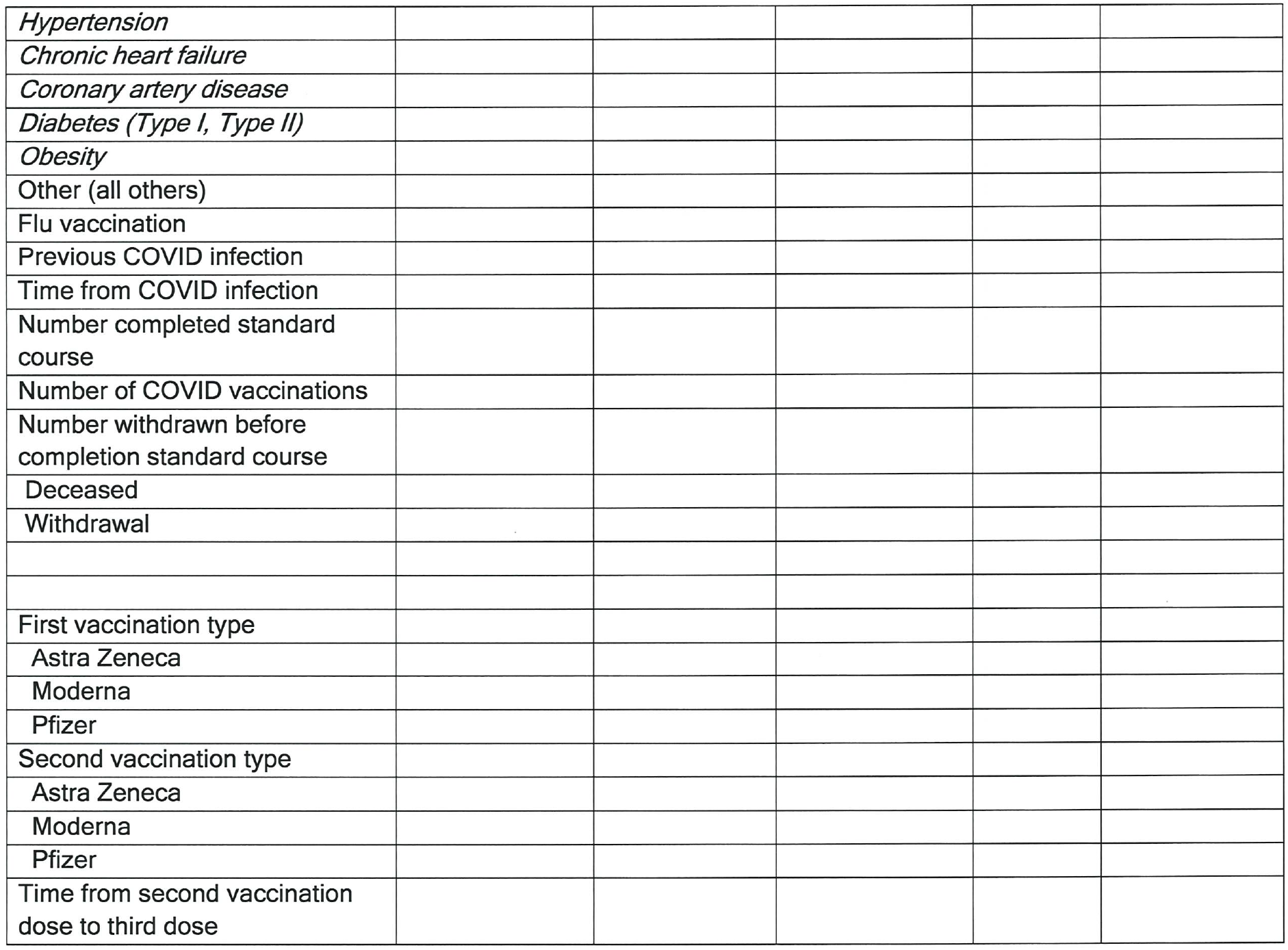
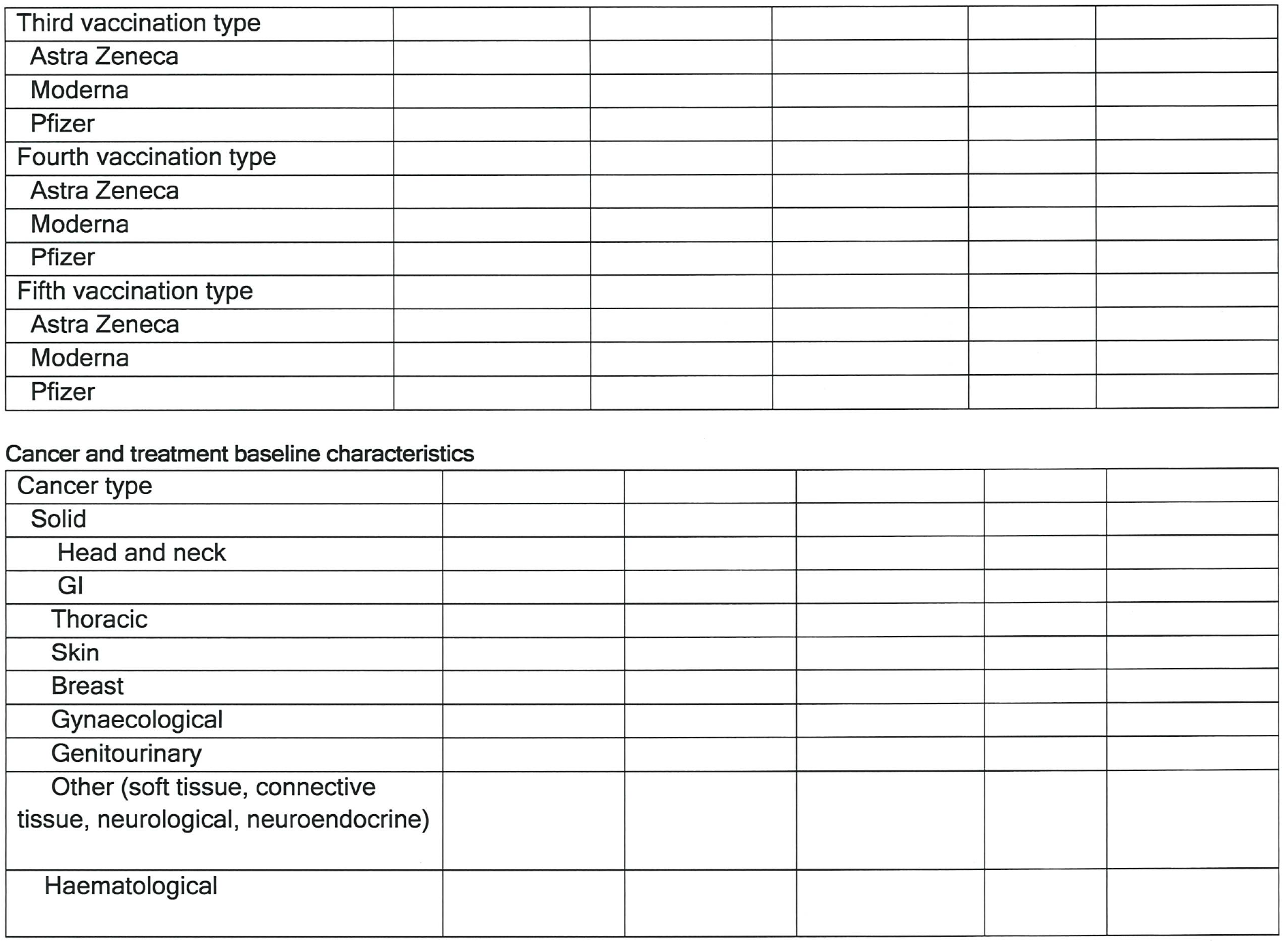
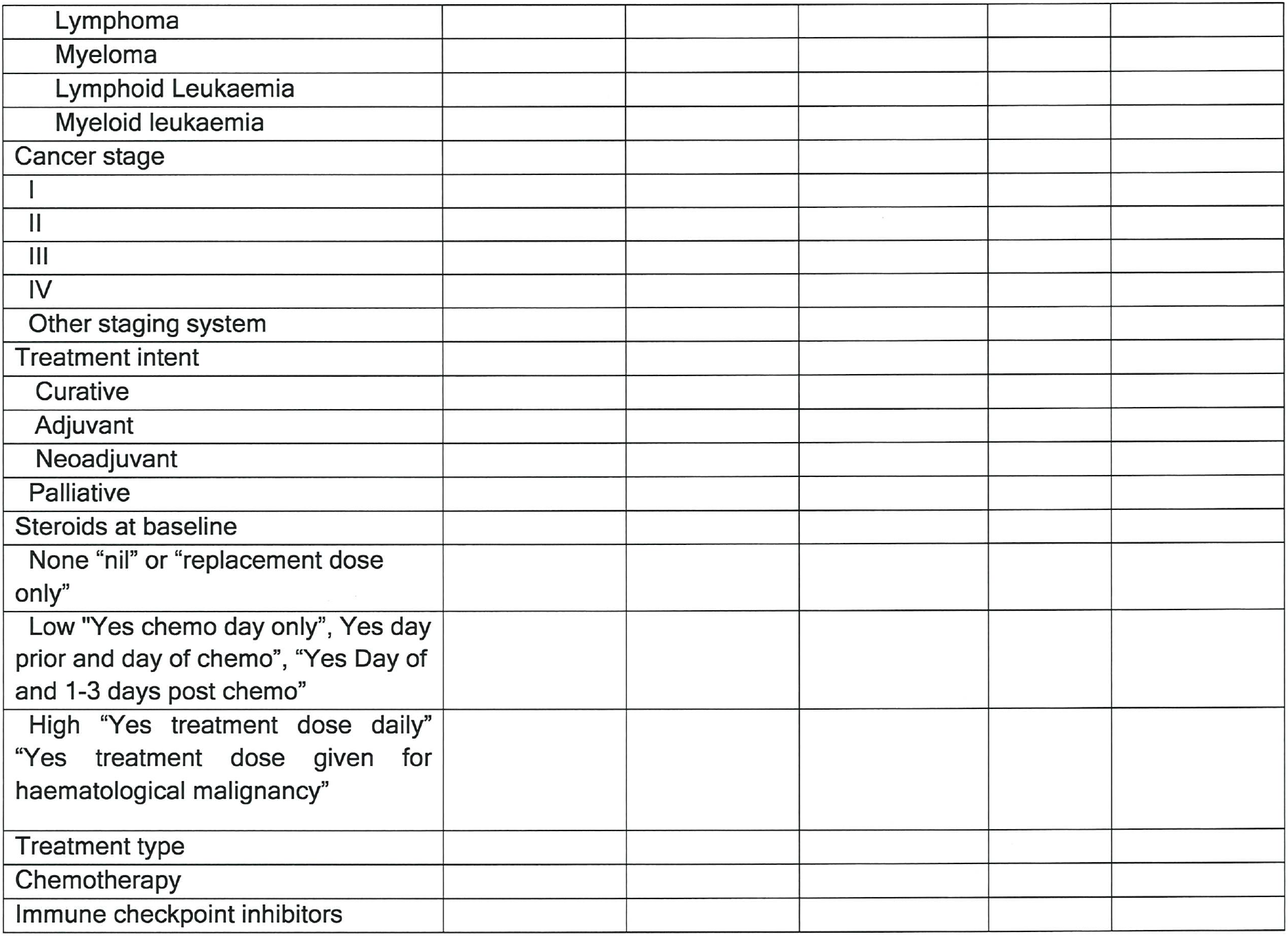
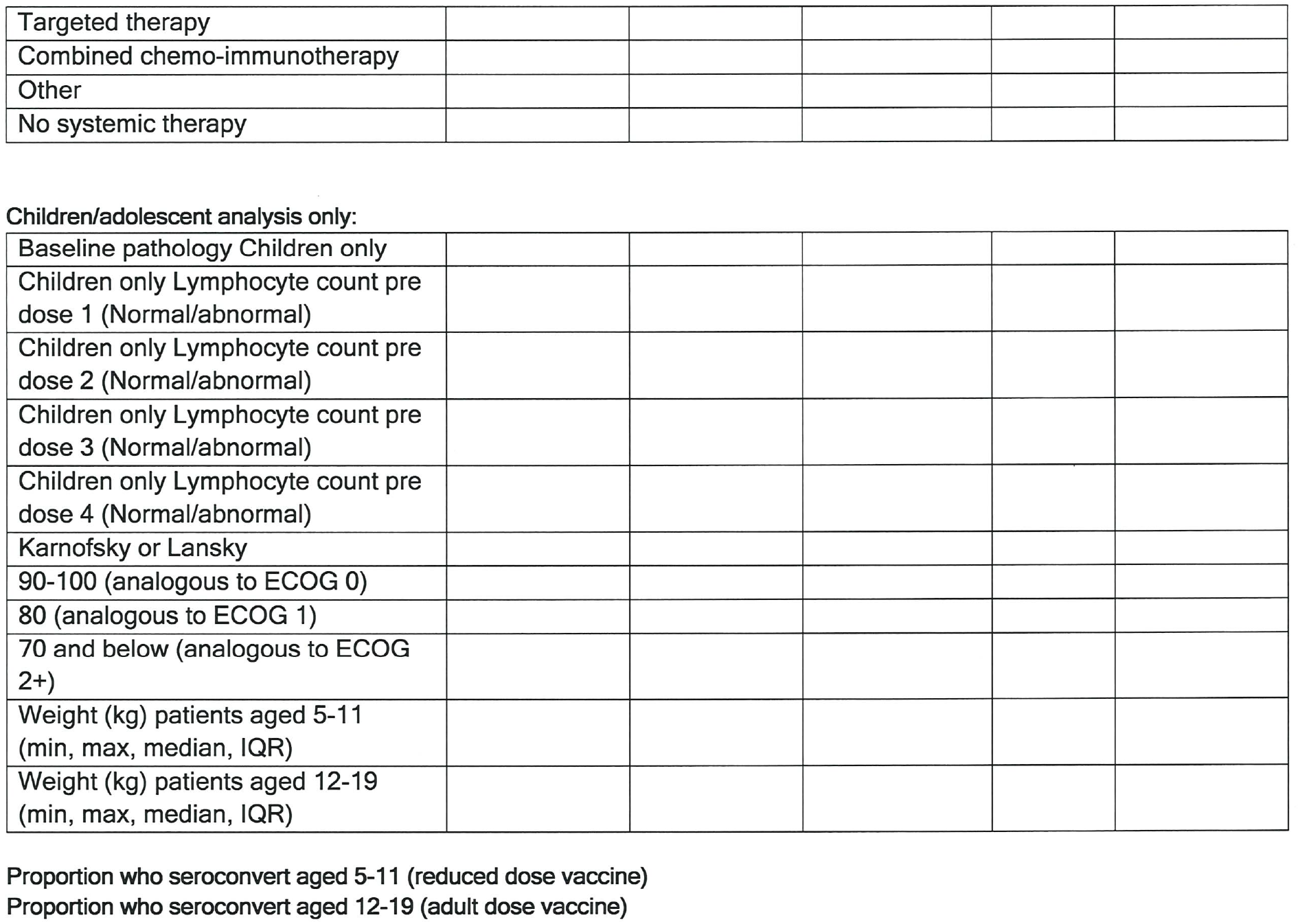
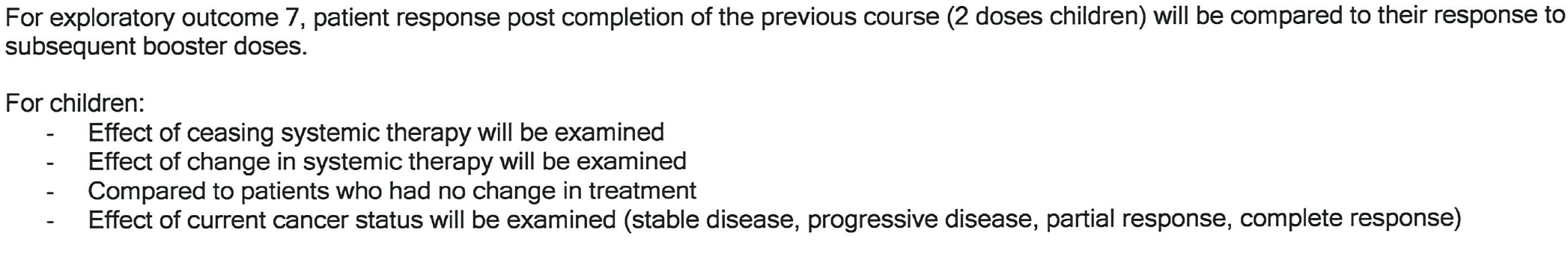
Participant baseline characteristics by cancer cohort

## Data Availability

All data produced in the present study are available upon reasonable request to the authors.

## List of Abbreviations

AE: Adverse Event
ATAGI: Australian Technical Advisory Group on Immunization
AZ: Astra Zeneca
CALD: Culturally And Linguistically Diverse
CDE: Common Data Elements
eCRF: Electronic Case Report Form
HREC: Human Research Ethics Committee
ICH-GCP: International Conference on Harmonization Good Clinical Practice
IgG: Immunoglobulin G
LBA: Ligand Binding Assay
Nab: Neutralizing Antibody
QoL: Quality of Life
PedsQL: Pediatric Quality of Life Scale
PRO-CTCAE: Patient Reported Outcomes Common Terminology Criteria for Adverse Events
MOGA: Medical Oncology Group of Australia
PEG: Polyethylene glycol
SAP: Statistical Analysis Plan
SARS: CoV-2 Severe Acute Respiratory Syndrome Coronavirus Disease
SerOzNET: SARS-CoV-2 post-vaccine surveillance studies in Australian children and adults with cancer

